# Suicide risk with selective serotonin reuptake inhibitors and new-generation serotonergic-noradrenergic antidepressants in adults: a systematic review and meta-analysis of observational studies

**DOI:** 10.1101/2020.05.11.20098178

**Authors:** Michael P Hengartner, Simone Amendola, Jakob A Kaminski, Simone Kindler, Tom Bschor, Martin Plöderl

## Abstract

**Background:** There is ongoing controversy whether antidepressant use alters the suicide risk in adult routine-care patients with depression and other treatment indications. The aim of this study was thus to examine the suicide risk with antidepressants in observational studies, considering financial conflicts of interest (fCOI) and publication bias.

**Design:** Systematic review and meta-analysis.

**Main outcome measures:** Risk of suicide, suicide attempt and/or intentional self-harm.

**Data sources:** We searched MEDLINE, PsycINFO, Web of Science, PsycARTICLES and SCOPUS for case-control and cohort studies published 1990-2020.

**Eligibility criteria for study selection:** Cohort and case-control studies in adults (aged ≥18 years) with depression and any unspecified condition reporting suicide risk for patients exposed to selective serotonin reuptake inhibitors (SSRI) or new-generation serotonergic-noradrenergic antidepressants (SNA) relative to unexposed patients.

**Data extraction and analysis:** Effects were aggregated with a random-effects model and reported as relative risk estimates (RE) with 95%-confidence-intervals. We assessed heterogeneity via I^2^-statistics and publication bias via funnel-plot asymmetry and trim-and-fill method. Study fCOI was defined present when lead-authors’ professorship was industry-sponsored, they received payments from the industry, or when the study was industry-sponsored.

**Results:** We included 27 original studies in the meta-analysis; 19 on depression (including other affective and anxiety disorders) and 8 on any unspecified condition. Use of SSRI or SNA for depression was associated with increased suicide risk (comprising both suicide and suicide attempt), RE=1.29, 1.06-1.57. Risk estimates were significantly lower in studies with fCOI (Q=21.87, p<0.001) and the trim-and-fill method estimated that 12 studies were missing due to publication bias; the result with missing studies imputed was RE=1.61, 1.31-1.99. Use of SSRI or SNA for all conditions (including depression and any unspecified condition) was associated with increased suicide risk, RE=1.43, 1.21-1.68. Studies with fCOI reported significantly lower risk estimates (Q=34.19, p<0.001) and the trim-fill method estimated that 13 studies were missing; after imputation of missing studies the result was RE=1.72, 1.44-2.05. Quality of evidence was rated very low due to substantial inconsistency of between-study results (I^2^≥85%).

**Conclusions:** Exposure to new-generation antidepressants is associated with increased suicide risk in adult routine-care patients with depression and other conditions. Publication bias and fCOI contribute to systematic underestimation of risk estimates in the published literature.

**Registration:** Open Science Framework, https://osf.io/eaqwn/

**Research in context:** *What is already known on this topic:* Meta-analyses of randomised controlled trials (RCT) found either no reduction of suicidal events or increased risk with new-generation antidepressants relative to placebo. However, generalisability of RCT findings is limited due to narrowly preselected participants, short study duration and treatment settings that are not representative of primary care routine practice. A previous meta-analysis of observational studies reported significantly reduced suicide risk in adults with depression exposed to selective serotonin reuptake inhibitors (SSRI), but it did not include new-generation serotonergic-noradrenergic antidepressants (SNA) and did not consider publication bias and study lead-authors’ financial conflicts of interest (fCOI).

*What this study adds:* Our results indicate that SSRI and SNA are associated with increased suicide risk in adult routine-care patients with depression and other psychiatric and non-psychiatric treatment indications. We further found empirical evidence for publication bias. Several studies with evidence of increased suicide risk with new-generation antidepressants likely remain unpublished. Accordingly, we found that study lead-authors with fCOI report significantly lower risk estimates than lead-authors without fCOI.

## Introduction

There is a longstanding and unresolved controversy whether selective serotonin reuptake inhibitors (SSRI) and new-generation serotonergic-noradrenergic antidepressants (SNA) such as duloxetine, venlafaxine, or mirtazapine alter the risk of suicide and suicide attempt in adults.^1-4^ While some suicidologists recommend antidepressants as an important suicide-prevention strategy,^5^ others argue that this strategy may increase the risk of suicide.^6^

The studies with the highest level of evidence - randomised controlled trials (RCT) - produced inconsistent findings. Some meta-analyses of RCT for the acute treatment of depression and other treatment indications in adults found significantly increased risk of suicidal events with antidepressants relative to placebo,^7-12^ but others did not.^13-17^ Two meta-analyses of long-term RCT have been conducted; both indicate that antidepressants may increase the risk of (attempted) suicide in patients with depression.^18 19^ Remarkably, no meta-analysis of RCT found a decreased suicide risk with antidepressants in comparison to placebo.

However, the findings from meta-analyses of RCT for depression have several caveats. A major limitation are the extensive exclusion criteria, for example substance abuse, comorbid disorders and acute suicidality.^20^ Participants included in RCT are therefore not representative of the average patient seen in routine practice.^21 22^ The experimental treatment setting of RCT, with its short study duration usually limited to a few weeks, its comprehensive assessments, close monitoring and frequent visits is neither representative of primary care routine practice, where most antidepressants are prescribed.^23 24^ Finally, systematic biases in the reporting of serious adverse events such as suicides and suicide attempts in favour of antidepressants appear to be pervasive and frequent in RCT.^14 25-27^

Well-designed observational studies with representative patients treated in routine practice may thus provide ecological validity. The last meta-analysis of observational studies was published in 2009 by Barbui et al.^28^ and it found that SSRI use relates to reduced suicide risk in adults with depression. Despite being published more than a decade ago, these findings are still influential in contemporary research, clinical guidance and scientific debate. For instance, based on the Barbui study a recent meta-review concluded that there is “highly suggestive” evidence that antidepressants protect against suicide in adults.^29^The aim of this study was thus to update the meta-analysis by Barbui et al.^28^ and to examine, whether incorporation of more recent evidence would support the conclusion that SSRI protect against suicide in adults with depression. Moreover, we expanded the Barbui study by focusing not only on SSRI, but also on SNA, and by including studies on depression and any psychiatric and non-psychiatric treatment indication. Given the substantial impact of selective publication and financial conflicts of interest (fCOI) in antidepressant research,^30-33^ we also attempted to control for these important biases.

## Methods

### Search strategy and selection criteria

We conducted our systematic review according to the PRISMA guideline.^34^ The review was registered with PROSPERO on January 21, 2020, but as of April 29, the submission was still being processed. As we were immediately informed that the evaluation process would take several months, the study protocol was also registered with the Open Science Framework on February 3, 2020 (https://osf.io/eaqwn/). We searched the databases MEDLINE, PsycINFO, Web of Science, PsycARTICLES and SCOPUS for all original studies in English published from 1990 to January 2020, using the terms “antidepressants”, “selective serotonin reuptake inhibitor”, “serotonin norepinephrine reuptake inhibitor”, and all individual new-generation antidepressants in combination with “suicide”, “suicide attempt”, or “self-harm”. The detailed search strategy and the Pubmed search term are shown in the appendix.

We additionally searched MEDLINE, PsycINFO and the German database PSYNDEX for non-English records, but found no additional studies that met our inclusion criteria. In order to find unpublished studies, we searched observational studies registered with ClinicalTrials.gov. One study met our inclusion criteria, but it was already included through our database search. Screening of titles and abstracts and subsequently assessments of full-texts were conducted independently by two investigators (SA and SK) during February and March 2020. Any discrepancies were resolved by consensus and, if necessary, through arbitration by the lead investigator (MPH).

The eligibility criteria were case-control and cohort studies in adults with depression or any other psychiatric and non-psychiatric treatment indication. We also included studies reporting risk estimates for any unspecified antidepressant class when it was possible to infer from the data or external sources (e.g. prescription rates for different antidepressant classes in the underlying population) that at least 75% of all prescriptions in the study sample were SSRI or SNA. We included only studies that report a risk estimate for an exposed group in relation to an unexposed group. Therefore, we excluded studies that compared the suicide risk with SSRI or SNA to other drugs (e.g. tricyclics) and studies that examined variations in suicide risk across different treatment phases within exposed subjects (e.g. suicide risk during acute therapy vs. continuation or maintenance therapy). In order to minimize confounding by indication, we required exposed and unexposed groups to be broadly comparable in terms of clinical and socioeconomic characteristics. We thus excluded studies that compared the suicide risk in antidepressant users with increased baseline suicide risk (e.g. psychiatric inpatients) to an unexposed group with low baseline risk (e.g. healthy people from the general population) if differences in baseline risk where statistically not controlled for (e.g. via propensity score matching or covariate adjustment). If the unexposed group consisted of patients with a baseline suicide risk comparable to the exposed group (e.g. when both groups comprised psychiatric inpatients), then we also included studies with unadjusted risk estimates.

### Outcomes

Our primary outcomes were the risk of suicide and suicide attempt in people exposed to new-generation antidepressants relative to an unexposed group according to the statistically best-adjusted analysis reported in the primary study. Suicide and suicide attempt were analysed separately and, to allow for comparability with the Barbui study,^28^ also as a composite outcome. We further conducted separate analyses for depression and any treatment indication unspecified, as well as for SSRI, SNA, and any new-generation antidepressant (comprising SSRI, SNA and any class unspecified). If studies reported not an overall risk estimate for the entire observation period, but instead separate estimates for mutually exclusive treatment periods (e.g. acute therapy, continuation and maintenance therapy), we always chose the risk estimate for the earliest treatment period. This decision was specified a priori in the protocol to minimise survivorship bias, as a consistent body of evidence indicates that the suicide risk is highest at the beginning of an illness episode when patients are acutely distressed.^35-38^

### Data analysis

To allow for comparability, study quality was rated with the same 10-point scale applied by Barbui et al.^28^ This scale assesses study quality based on six domains (population framework; study design; description of demographic data; description of clinical data; description of outcome data; and covariate adjustment) and ranges from 0 (minimal quality) to 10 (maximal quality) points.^39 40^ The scale and its rating system are shown in the appendix. The quality of evidence for each outcome was evaluated according to the GRADE system.^41 42^ Study fCOI was coded present when a lead-author (i.e. first or last author) had received payments from the pharmaceutical industry (e.g. as speaker, consultant or adviser); when the professorship of a lead-author was funded by the pharmaceutical industry; or when the study was sponsored by the pharmaceutical industry. For it we not only assessed the fCOI statement made in the primary study, but also searched the internet and declarations in other publications by the same authors.

We used an inverse variance random-effects model and computed relative risk estimates (RE) with 95% confidence intervals based on the odds ratios or hazard ratios extracted from the primary studies. We used the Sidik-Jonkman and Hartung-Knapp adjustments recommended in the Cochrane Handbook.^43^ Publication bias was inferred from funnel-plot asymmetry and tested with the trim-and-fill method.^44^ Heterogeneity (inconsistency of between-study results) was estimated with I^2^-statistics. All analyses were conducted with the R packages metafor and metagen. The R-code and raw data are available online, https://osf.io/eaqwn/.

### Patient involvement

Patients or the public were not involved in the design, or conduct, or reporting, or dissemination plans of our research.

## Results

### Search results and inclusion of primary studies

The literature search identified 9542 records, which were reduced to 6676 after removal of duplicates. After screening of titles and abstracts, further 6602 records were excluded and 74 full-text were assessed for eligibility. A final number of 32 studies were retained. Of these, 5 focused on unique conditions other than depression, other affective disorders or anxiety disorders. These studies were not included in the quantitative synthesis (meta-analysis) but the results are reported qualitatively in the appendix. Altogether 27 studies, of which 19 on depression (plus other affective disorders and anxiety disorders) and 8 on any treatment indication unspecified were included in the meta-analysis. Included in this dataset are also the 6 adult studies incorporated in the Barbui meta-analysis.^28^ The study selection process is shown in Figure 1 and the characteristics of the primary studies included in the meta-analysis are summarized in the appendix. Briefly, 8 studies were conducted in North America (7 in the USA and 1 in Canada), 17 studies in Europe and 2 elsewhere (1 in Australia and 1 in New Zealand). 9 studies had fCOI (among them all 6 studies included the Barbui study) and 18 had no fCOI. Study quality ratings ranged from 4 to 9 points (median=8, interquartile range=2). Detailed quality ratings for all studies are shown in the appendix.

**Figure 1:**
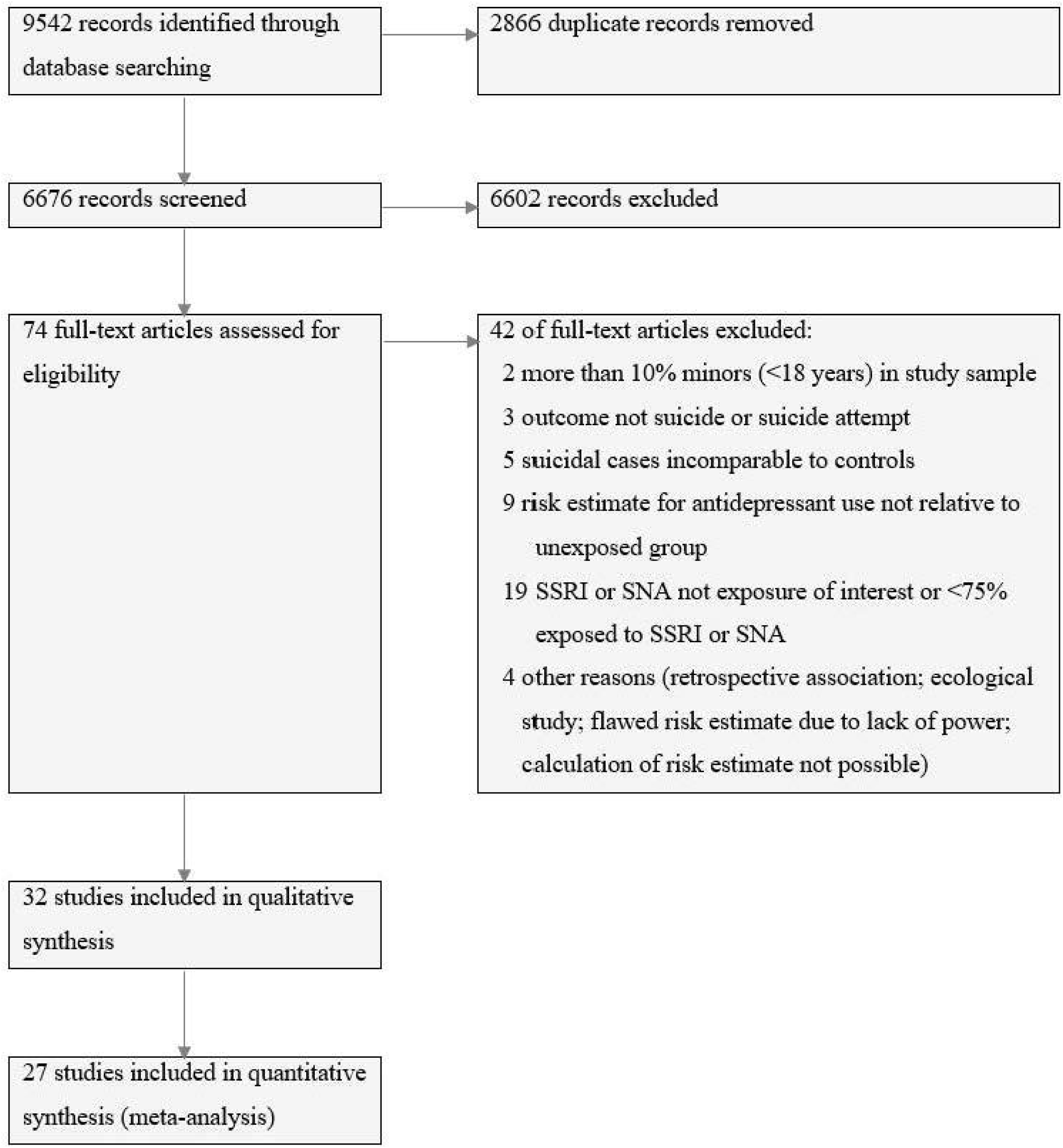
PRISMA flow chart

### Meta-analyses for depression

The meta-analytic results for the association between exposure to new-generation antidepressants and suicide risk in depression studies are shown in Figure 2. The summary effect estimate for suicide with any new-generation antidepressant was RE=1.29, 0.90-1.86. For SSRI the effect estimate was RE=1.09, 0.51-2.33 and for SNA it was RE=1.31, 0.62-2.76. The funnel-plot was asymmetrical and the trim-and-fill method suggested 5 missing studies. Imputation of missing studies revealed a significantly increased risk, RE=1.64, 1.14-2.36. Funnel-plots for all outcomes and Egger’s test for funnel-plot asymmetry are shown in the appendix. Study fCOI was a significant moderator (Q=6.35, df=1, p=0.012); studies with fCOI (RE=1.00, 0.75-1.34) reported considerably lower risk estimates than studies without fCOI (RE=2.29, 0.99-5.28). Forest-plots for fCOI subgroups are shown in the appendix.

**Figure 2:**
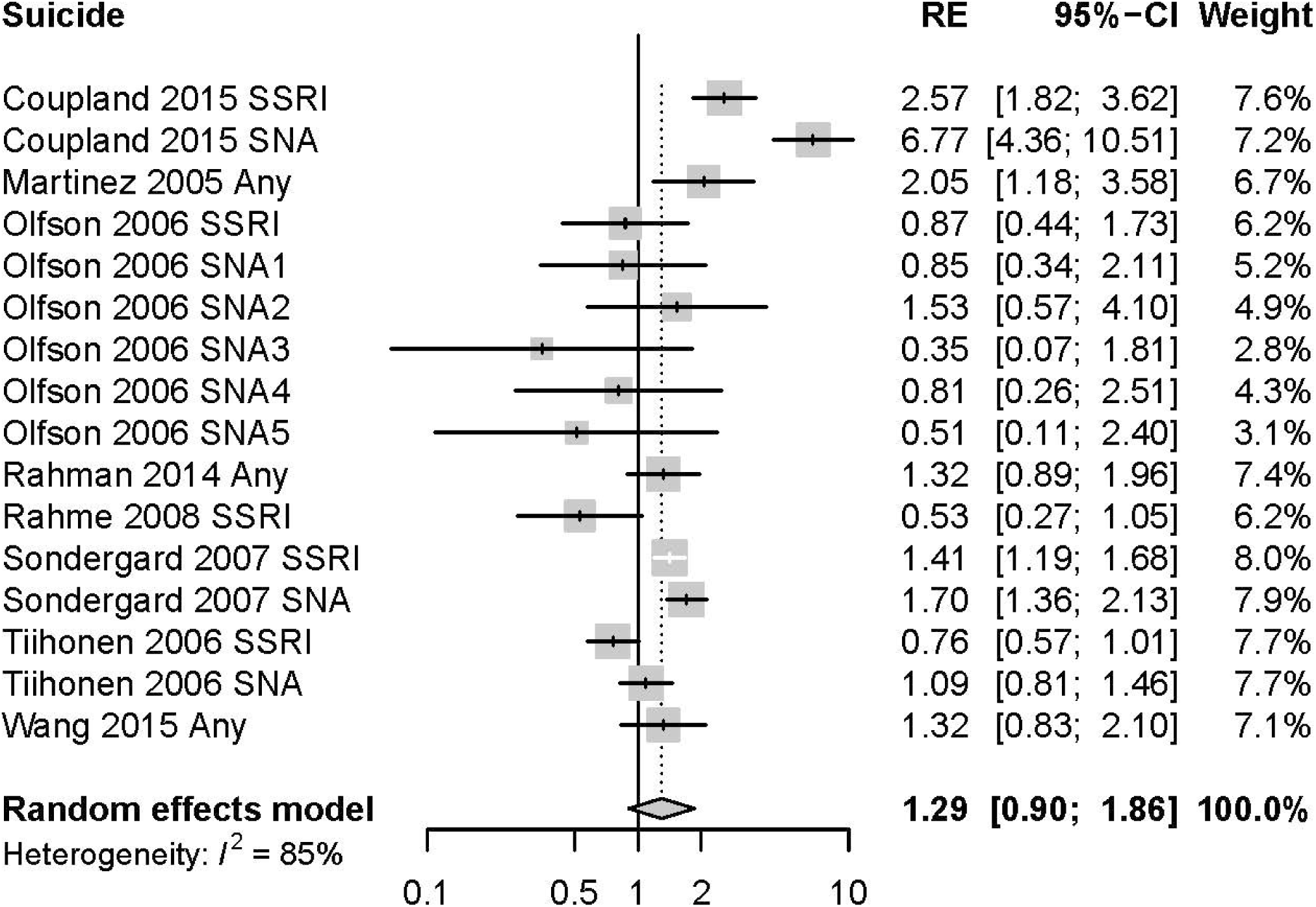
Forest-plots for suicide risk in patients with depression (including other affective disorders and anxiety disorders). SSRI: selective serotonin reuptake inhibitor; SNA: new-generation serotonergic-noradrenergic antidepressant (includes serotonin norepinephrine reuptake inhibitor, e.g. duloxetine and venlafaxine, and atypical antidepressants, e.g. mirtazapine and bupropion); S: Suicide; SA: Suicide attempts; RE: risk estimate

For suicide attempt the summary effect estimate for any new-generation antidepressant was RE=1.31, 1.03-1.68 (for SSRI, RE=0.99, 0.57-1.72; for SNA, RE=1.32, 0.79-2.23). Trim-and-fill method estimated that 8 studies were missing and with imputation of these studies the result was RE=1.61, 1.24-2.10. Study fCOI had a significant effect (Q=12.86, df=1, p<0.001); studies with fCOI (RE=0.91, 0.70-1.17) reported considerably lower risk estimates than studies without fCOI (RE=1.84, 1.31-2.58). The summary effect estimate for suicide and suicide attempt combined (composite outcome) with any new-generation antidepressant was RE=1.29, 1.06-1.57 (for SSRI, RE=1.03, 0.70-1.51; for SNA, RE=1.28, 0.86-1.90). Trim-and-fill method suggested that 12 studies were missing and the result with imputation of missing studies was RE=1.59, 1.28-1.98. Study fCOI was again a significant moderator (Q=21.87, df=1, p<0.001); studies with fCOI showed no clear association (RE=0.91, 0.77-1.09), whereas studies without fCOI reported significantly increased suicide risk (RE=1.94, 1.46-2.59). Of 12 studies that reported risk estimates for SSRI, 8 studies (67%) had fCOI and 4 studies (33%) had no fCOI. By contrast, among the 8 studies reporting risk estimates for any class unspecified, none (0%) had fCOI.

### Meta-analyses for all treatment indications

The meta-analytic results for all treatment indications are shown in Figure 3. The risk estimate for suicide with any new-generation antidepressant was RE=1.44, 1.15-1.80 (for SSRI, RE=1.33, 0.86-2.04; for SNA, RE=1.28, 0.74-2.21). Trim-and-fill method estimated that 5 studies were missing and the corresponding result after imputation of missing studies was RE=1.62, 1.28-2.06. Study fCOI had a significant effect (Q=11.36, df=1, p<0.001). Studies with fCOI reported no increased risk (RE=1.08, 0.88-1.33), but studies without fCOI did (RE=1.98, 1.43-2.76).

**Figure 3:**
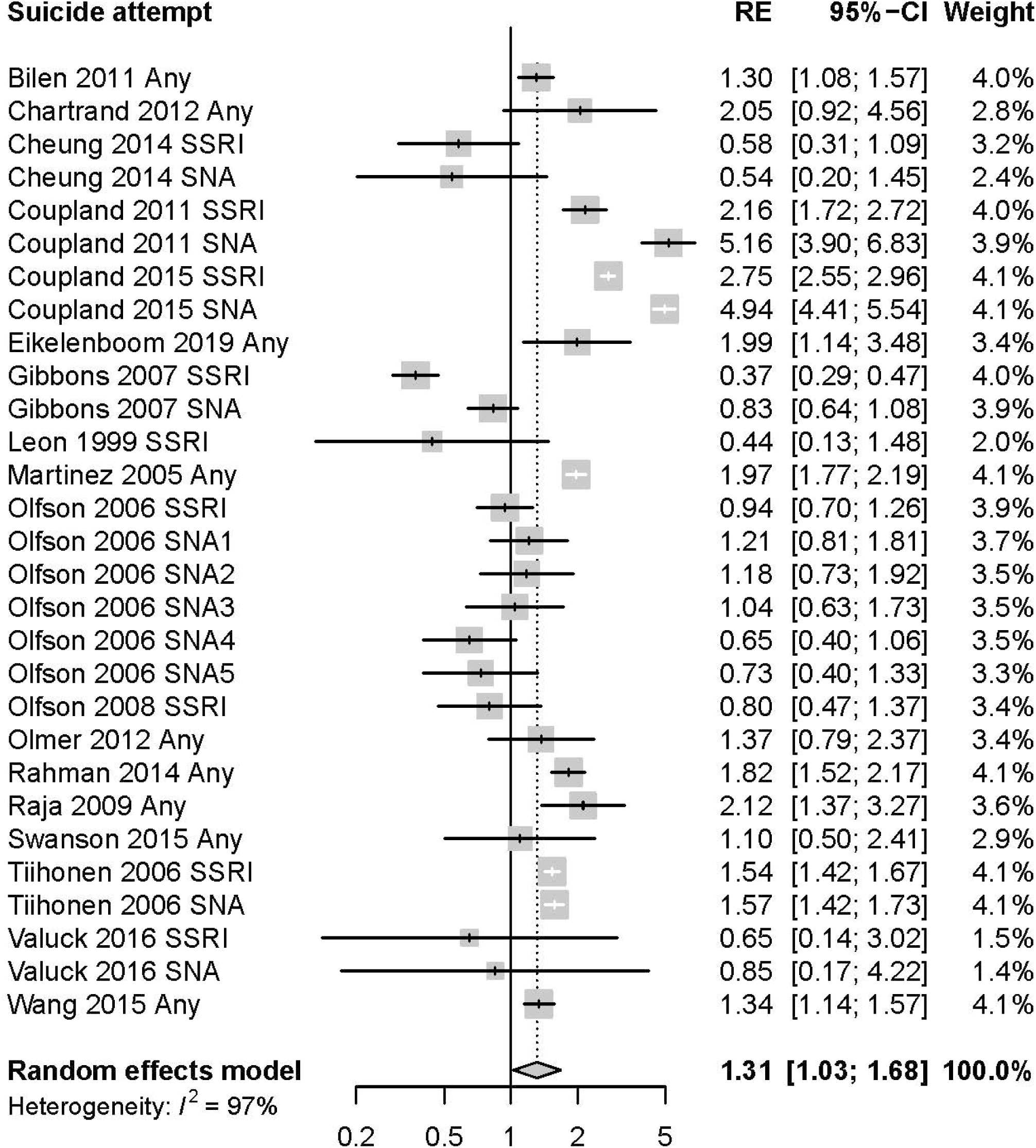
Forest-plots for suicide risk in patients with any treatment indication (including depression and any unspecified psychiatric and non-psychiatric condition). SSRI: selective serotonin reuptake inhibitor; SNA: new-generation serotonergic-noradrenergic antidepressant (includes serotonin norepinephrine reuptake inhibitor, e.g. duloxetine and venlafaxine, and atypical antidepressants, e.g. mirtazapine and bupropion); S: Suicide; SA: Suicide attempts; RE: risk estimate

The risk of suicide attempt with any new-generation antidepressant was RE=1.47, 1.17-1.85 (for SSRI, RE=1.08, 0.67-1.75; for SNA, RE=1.34, 0.81-2.21). Trim-and-fill method suggested that 7 studies were missing and the result after imputation of missing studies was RE=1.79, 1.40-2.28. Study fCOI was a significant moderator (Q=21.97, df=1, p<0.001); no clear effect was shown in studies with fCOI (RE=0.91, 0.70-1.17), but studies without fCOI revealed significantly increased risk (RE=2.05, 1.57-2.67).

The risk of suicide and suicide attempt combined with any new-generation antidepressant was RE=1.45, 1.23-1.70 (for SSRI, RE=1.19, 0.88-1.60; for SNA, RE=1.28, 0.90-1.80). Trim-and-fill method estimated that 14 studies were missing; the result after imputation of missing studies was RE=1.75, 1.47-2.08. Likewise, there was a significant effect for fCOI (Q=37.17, df=1, p<0.001); studies with fCOI reported no increased risk (RE=0.96, 0.82-1.12), but studies without fCOI showed significantly increased risk (RE=2.02, 1.66-2.46).

The quality of evidence according to GRADE was rated very low for all outcomes due to substantial inconsistency of between-study results (I^2^≥85%, see Figures 2 and 3).

### Sensitivity and subgroup analysis

Three effect estimates were found to be potential outliers. A sensitivity analysis with these potential outlier effects excluded revealed no meaningful differences compared to the results based on the full dataset (see supplementary material). The subgroup analyses for the composite outcome in association with any new-generation antidepressant are reported in Table 1. No significant subgroup differences were found for main treatment indication (depression vs. any unspecified), age (old adults vs. young adults), study design (cohort vs. case-control), study quality (high vs. low), and covariate adjustment (yes vs. no). There was a significant subgroup-difference for antidepressant class. Risk estimates for SSRI (RE=1.19, 0.88-1.60) and SNA (RE=1.28, 0.90-1.80) did not differ meaningfully, but studies that examined any class unspecified reported higher risk estimates (RE=1.87, 1.55-2.25). North American studies (all but one from the USA) reported significantly *reduced* suicide risk with new-generation antidepressants (RE=0.82, 0.68-0.99), whereas European studies showed significantly *increased* risk (RE=1.82, 1.51-2.20).

**Table 1:** Risk of suicide and suicide attempt combined in association with exposure to new-generation antidepressants

|  |  | Number of comparisons | Summary estimate | 95% Confidence interval |  | Heterogeneity | Subgroup difference |
| --- | --- | --- | --- | --- | --- | --- | --- |
|  |  | k | RE | CILB | CIUB | I<sup>2</sup> |  |
| Drug class | SSRI | 19 | 1.19 | 0.88 | 1.60 | 96.0 | Q = 9.39<br>p = 0.009 |
|  | SNA | 21 | 1.28 | 0.90 | 1.80 | 96.2 |  |
|  | Any unspecified | 21 | 1.87 | 1.55 | 2.25 | 89.3 |  |
| Main treatment indication | Depression | 41 | 1.35 | 1.10 | 1.65 | 95.6 | Q = 1.53<br>p = 0.217 |
|  | Any unspecified | 20 | 1.65 | 1.26 | 2.17 | 93.1 |  |
| Location | North America | 21 | 0.82 | 0.68 | 0.99 | 65.5 | Q = 39.95<br>p < 0.001 |
|  | Europe | 36 | 1.82 | 1.51 | 2.20 | 95.7 |  |
|  | Other | 4 | 1.73 | 0.84 | 3.57 | 10.8 |  |
| Older adult sample (≥65 years) | Yes | 4 | 1.67 | 0.35 | 7.86 | 94.7 | Q = 0.09<br>p = 0.764 |
|  | No | 54 | 1.44 | 1.21 | 1.71 | 95.0 |  |
|  | Missing data | 3 |  |  |  |  |  |
| Study Design | Cohort | 37 | 1.59 | 1.27 | 1.99 | 96.4 | Q = 3.16<br>p = 0.076 |
|  | Case-control | 24 | 1.22 | 1.00 | 1.50 | 79.7 |  |
| Study quality | High (≥7 points) | 49 | 1.36 | 1.15 | 1.61 | 94.4 | Q = 2.38<br>p = 0.123 |
|  | Low (<7 points) | 12 | 1.92 | 1.21 | 3.03 | 96.5 |  |
| Covariate adjustment | Yes | 51 | 1.40 | 1.19 | 1.65 | 94.1 | Q = 0.54<br>p = 0.460 |
|  | No | 10 | 1.71 | 0.96 | 3.04 | 97.2 |  |
| fCOI | Yes | 28 | 0.96 | 0.82 | 1.12 | 86.1 | Q = 37.17<br>p < 0.001 |
|  | No | 33 | 2.02 | 1.66 | 2.46 | 94.3 |  |

SSRI: selective serotonin reuptake inhibitors; SNA: new-generation serotonergic-noradrenergic
antidepressants (includes serotonin norepinephrine reuptake inhibitors, e.g. duloxetine and
venlafaxine, and atypical antidepressants, e.g. mirtazapine and bupropion); fCOI: financial conflicts of
interest; RE: risk estimate; CILB: confidence interval lower bound; CIUB: confidence interval upper
bound

The fCOI results for all outcomes are reported in detail above and the subgroup forest-plot is shown in Figure 4. In studies without fCOI visual inspection of the funnel-plot indicated no asymmetry. This finding was statistically supported by Egger’s test, t=-0.8456, df=31, p=0.404. By contrast, in studies with fCOI the funnel-plot was asymmetrical, both visually and statistically, t=-2.1368, df=26, p=0.042. Both funnel-plots are shown in the appendix. Study fCOI and study location were related; while 6 of 8 (75%) North American studies had fCOI, only 3 of 17 (18%) European studies had so. Study fCOI was also related to drug class examined. While 9 of 16 studies (56%) on SSRI or SNA had fCOI, 0 of 11 studies (0%) on any class unspecified had so. The significant subgroup differences for drug class and study location are thus at least in part attributable to study fCOI.

**Figure 4:**
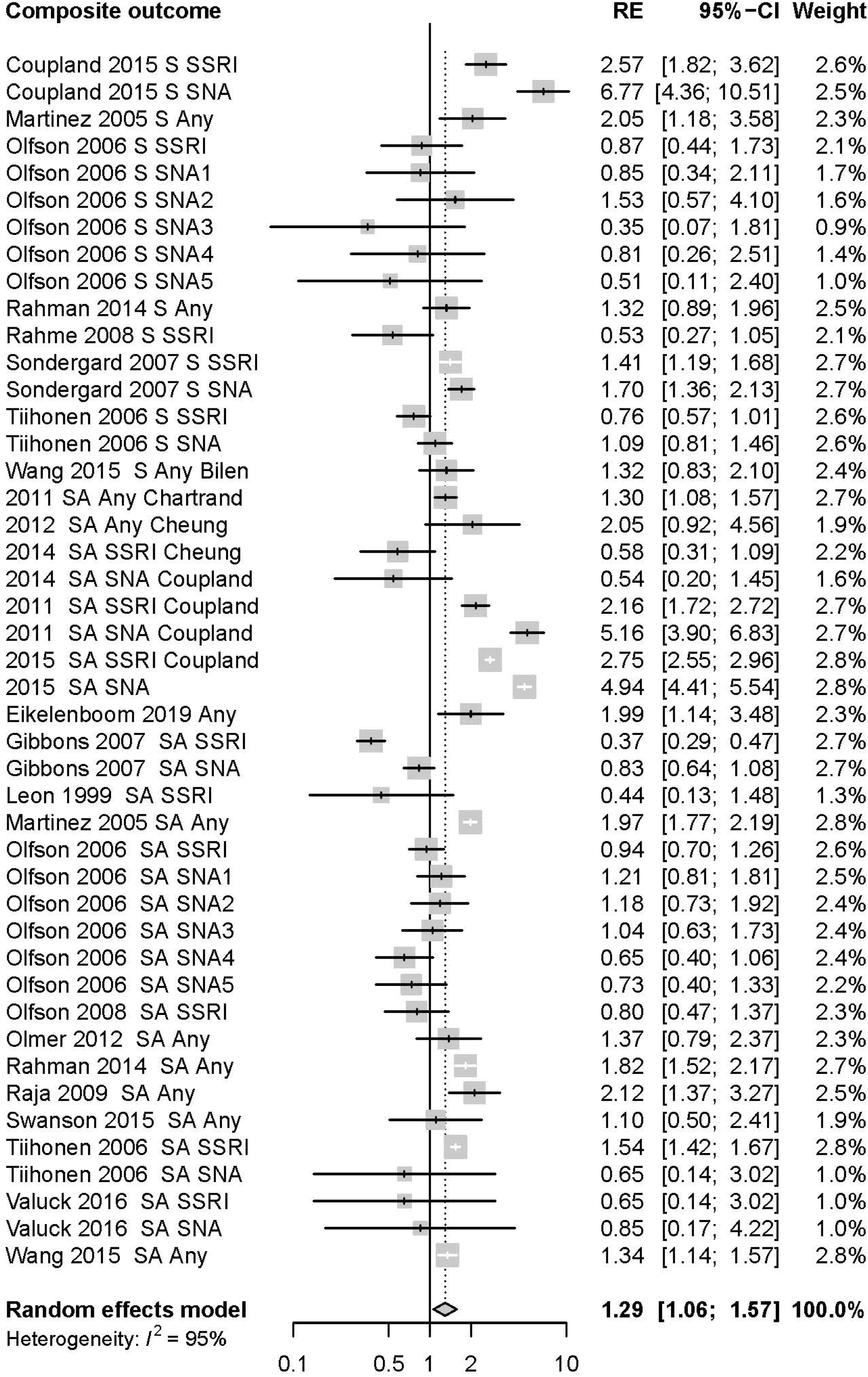
Forest-plot for suicide risk by financial conflicts of interest (fCOI) subgroups. SSRI: selective serotonin reuptake inhibitor; SNA: new-generation serotonergic-noradrenergic antidepressant (includes serotonin norepinephrine reuptake inhibitor, e.g. duloxetine and venlafaxine, and atypical antidepressants, e.g. mirtazapine and bupropion); S: Suicide; SA: Suicide attempts; RE: risk estimate

## Discussion

Our meta-analysis of observational studies revealed no clear association between exposure to SSRI and suicide risk in patients with depression and therefore contradicts the reduced suicide risk reported in the previous meta-analysis by Barbui and colleagues from 2009.^28^ While the Barbui study relied on 9 comparisons from 6 primary studies, our updated meta-analysis included 14 comparisons from 11 primary studies (including all 6 primary studies from the Barbui meta-analysis plus 5 new studies). It is also important to note that all six adult studies on SSRI for depression included in Barbui et al.^28^ were conducted by lead-authors with fCOI. In view of the significant association between fCOI and lower risk estimates detected in our analysis, this has likely resulted in systematically underestimated risk estimates in the Barbui study. Moreover, our study revealed a significantly increased suicide risk with any new-generation antidepressant in patients with depression as well as in patients with any treatment indication unspecified. Our results are thus consistent with various meta-analyses of FDA safety summaries for acute depression trials that revealed an increased suicide risk with new-generation antidepressants as a group relative to placebo.^9, 10, 12, 45^ A significantly increased risk of suicidal events was also meta-analytically demonstrated with SSRI in general^8^ and specifically with paroxetine in meta-analyses of published and unpublished placebo-controlled short-term trials.^7 11^ Our results also correspond with two meta-analyses of long-term depression trials, which found an increased risk of (attempted) suicide with antidepressants.^18 19^

We rated the quality of evidence very low due to substantial inconsistency of between-study results. Effect estimates are thus very uncertain and may change substantially with publication of further studies. Moreover, there was evidence of publication bias according to asymmetrical funnel-plots. Smaller studies reporting reduced suicide risk with antidepressants were overrepresented, which suggests that various studies with increased risk estimates remain unpublished. After correcting for publication bias via trim-and-fill method, the summary effect estimates indicated a considerably increased suicide risk with new-generation antidepressants. These findings are consistent with the literature on selective publication of antidepressant trials^33 46^ and misreporting of serious adverse events in published antidepressant trials.^14 25^ These biases result in systematically inflated efficacy estimates and underestimation of harms.^31 47 48^ We further found that studies with fCOI reported significantly lower risk estimates than studies without fCOI. Moreover, publication bias according to asymmetrical funnel-plots was detected in studies with fCOI, but not in studies without fCOI. These findings are consistent with the literature showing that fCOI systematically bias the benefit-to-risk assessment of drugs in favour of the industry, both in psychiatry^32 49-51^ and in other medical fields.^52-55^ In our set of depression studies, two-thirds of studies focusing on SSRI had fCOI, whereas no study on any class unspecified had so. This may explain why in depression studies no increased risk was shown specifically for SSRI whereas significantly increased risk was found for any new-generation antidepressant.

### Limitations

Population-based register-linkage studies allow studying very large and representative samples and to control for various socioeconomic and clinical covariates, which increases generalisability of findings and which minimizes confounding by indication. Yet these studies may have important limitations, such as incomplete assessment of exposure periods. For instance, in the cohort study by Tiihonen et al.,^56^ the register used to assess antidepressant exposure contained only the purchase of medication from pharmacies in outpatient care, but not prescriptions from hospital care. This means that suicides occurring in hospital or shortly after discharge were necessarily ascribed to the unexposed period in many first-episode patients, although it is likely that in these cases antidepressants were prescribed by a hospital doctor. As the suicide risk is highest in the first weeks after hospital admission and immediately after discharge,^57^ this incomplete assessment of exposure may result in an underestimation of the suicide risk in the exposed group. On the other hand, prescription registries record only whether drugs had been dispensed, but not whether the drugs were actually taken as prescribed. It is known that in pharmacotherapy for chronic conditions patients show rather low adherence rates.^58^ If some people redeemed a prescription but did not use the drugs for any reason, this may result in an overestimation of the suicide risk in the exposed group.

Another issue is confounding by indication. Although most studies tried to minimize this bias by controlling for important covariates such as previous suicide attempts and depression severity, unlike RCT it cannot be excluded that patients with more severe forms of psychopathology were more likely to receive antidepressants. On the other hand, most SSRI and SNA are prescribed by GPs,^23 24^ often for mild and subclinical depression.^59 60^ Therefore, antidepressant use may not necessarily indicate more severe psychopathology in people with depression. For instance, Olfson et al.^24^ showed that US patients with serious psychological distress (a strong risk factor for suicide) are no more often treated with antidepressants than patients with less serious or no distress. This resonates with our meta-analytic finding that adjusted risk estimates were not significantly lower than unadjusted estimates. Remarkably, the two studies that reported the strongest suicide-protective effects were both unadjusted.^61 62^ Depending on the sample studied, confounding by indication could thus result in both over- and underestimation of risk estimates. In accordance, covariate adjustment was not able to explain the high inconsistency in between-study results. To better explain heterogeneity of risk estimates, more detailed methodological aspects presumably need to be considered (e.g. concomitant psychosocial support).

## Conclusions

Our systematic review and meta-analysis of observational studies revealed that use of new-generation antidepressants is associated with increased suicide risk in adult patients, especially after controlling for publication bias and fCOI. Many studies showing increased suicide risk with antidepressants likely remain unpublished. Relatedly, studies with fCOI report significantly lower risk estimates than studies without fCOI. This has two important implications. First, contrary to prominent claims^5 29^ we find no reliable evidence indicating that antidepressants protect against suicide. Instead, it appears that antidepressant use may even increase suicide risk. Associations detected in observational studies do not necessarily imply causality, but there is evidence of a causal relationship according to various meta-analyses of RCT^7 8 10 18 63^ and challenge-dechallenge-rechallenge testing protocols.^64-67^ Second, our findings underline the necessity of even more stringent research regulation to exclude possible bias through fCOI.^68-70^ In accordance with the literature, ^32 52 53 55^ our results suggest that studies with fCOI are systematically biased. Therefore, public access to raw data and independent research on these data conducted by authors without fCOI is warranted.

## Data Availability

Data and statistical code will be made freely available upon publication of the study in a scientific journal

https://osf.io/eaqwn/

## Funding

JAK is supported by the Charité Clinician-Scientist Program of the Berlin Institute of Health. SA is supported by the Scholarships for Advanced Studies Abroad of the Sapienza University of Rome. The sponsors had no influence on the conduct of this study. No funding was received for this specific study.

## Declaration of interests

All authors have completed the ICMJE uniform disclosure form at http://www.icmje.org/coi disclosure.pdf and declare: no support from any organisation for the submitted work; no financial relationships with any organisations that might have an interest in the submitted work in the previous three years, no other relationships or activities that could appear to have influenced the submitted work.

---

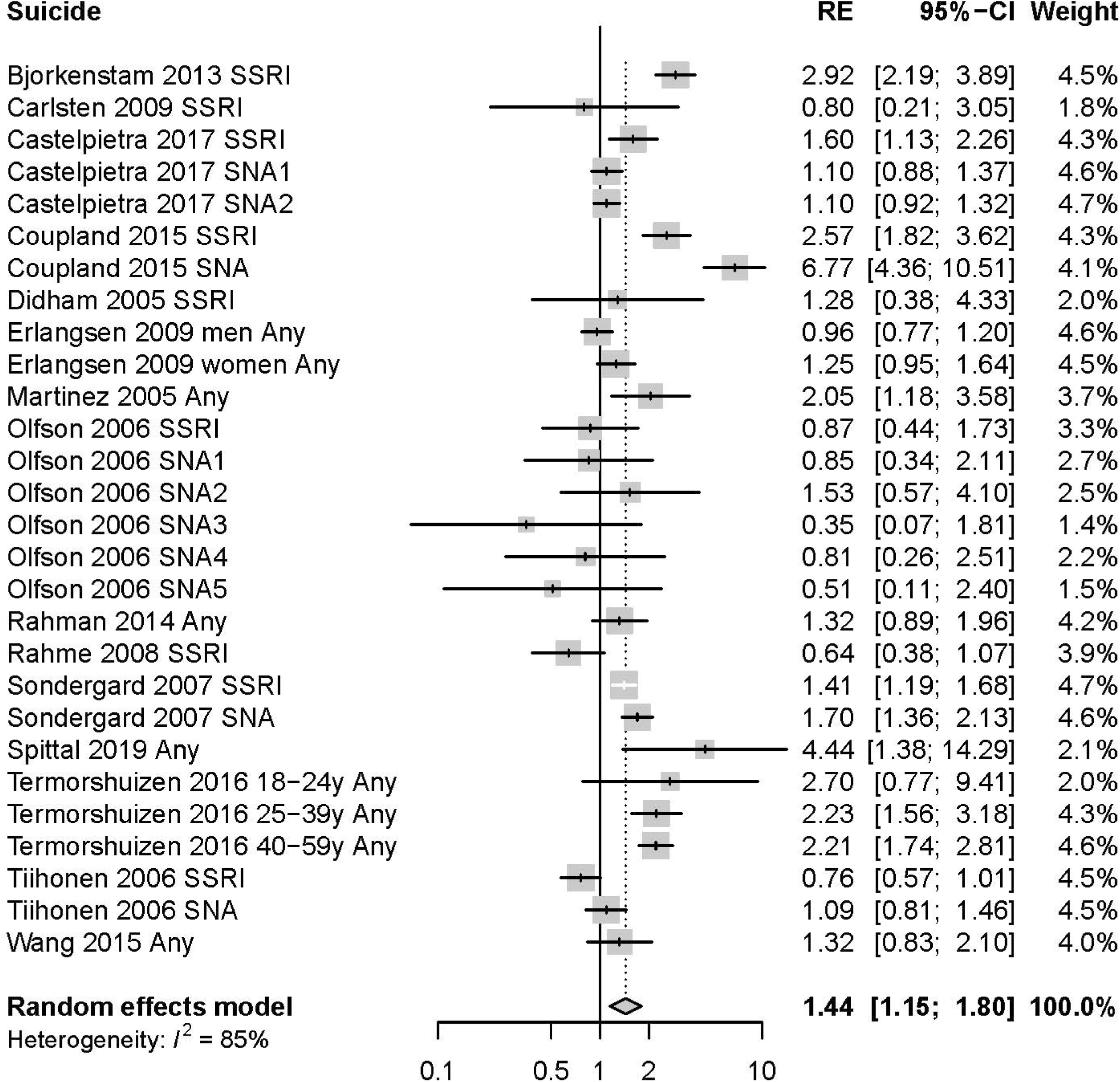

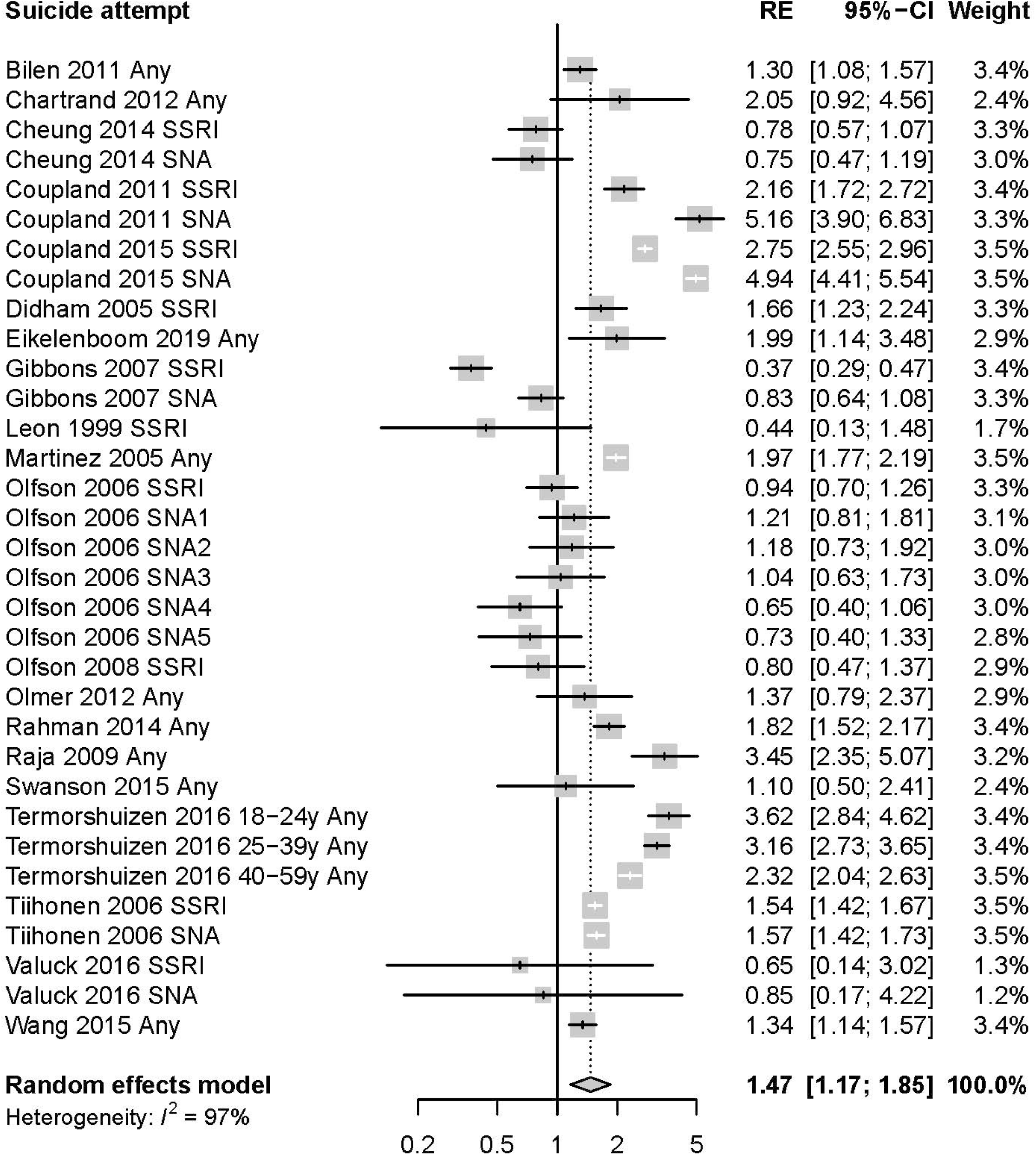

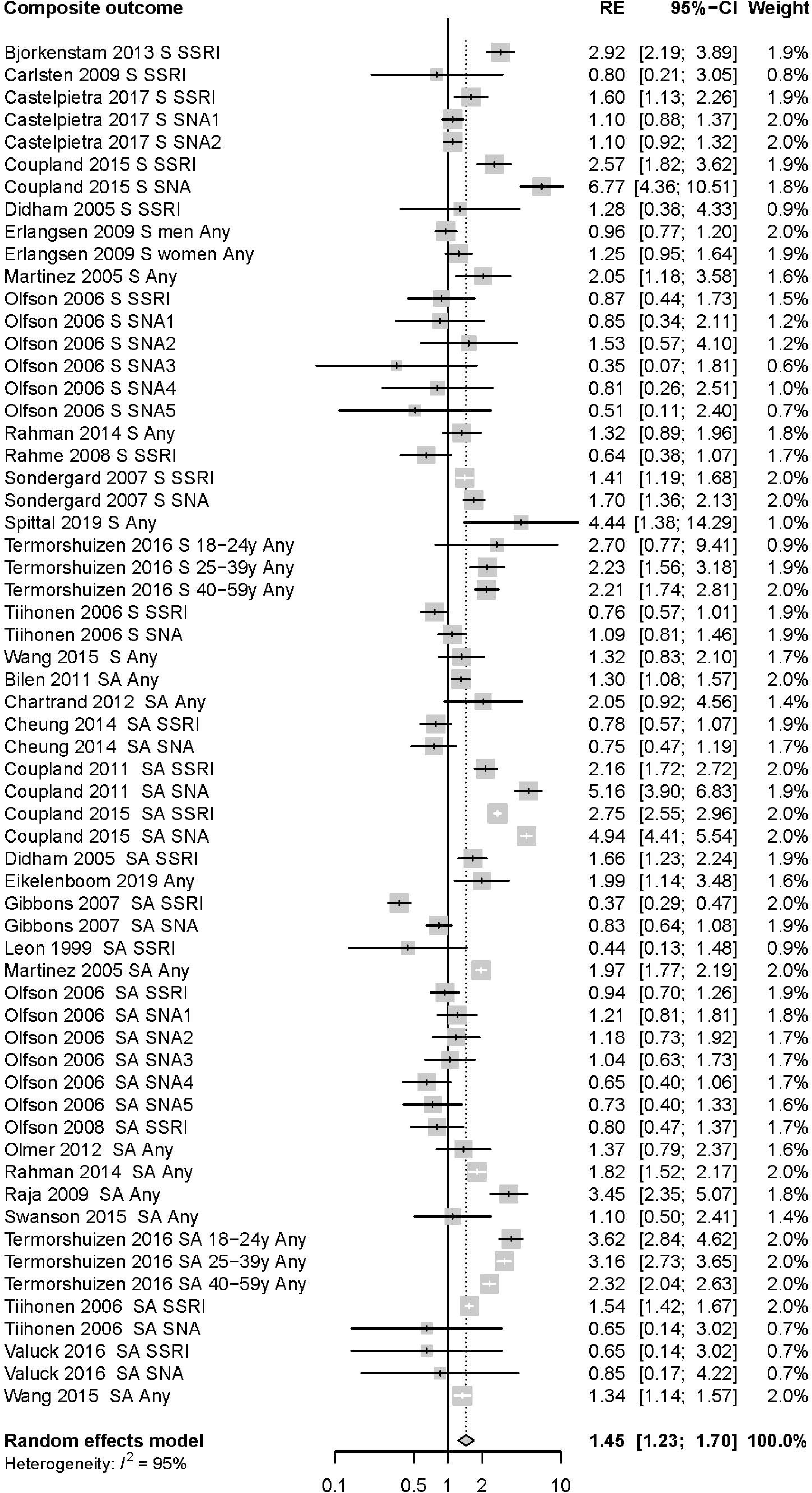

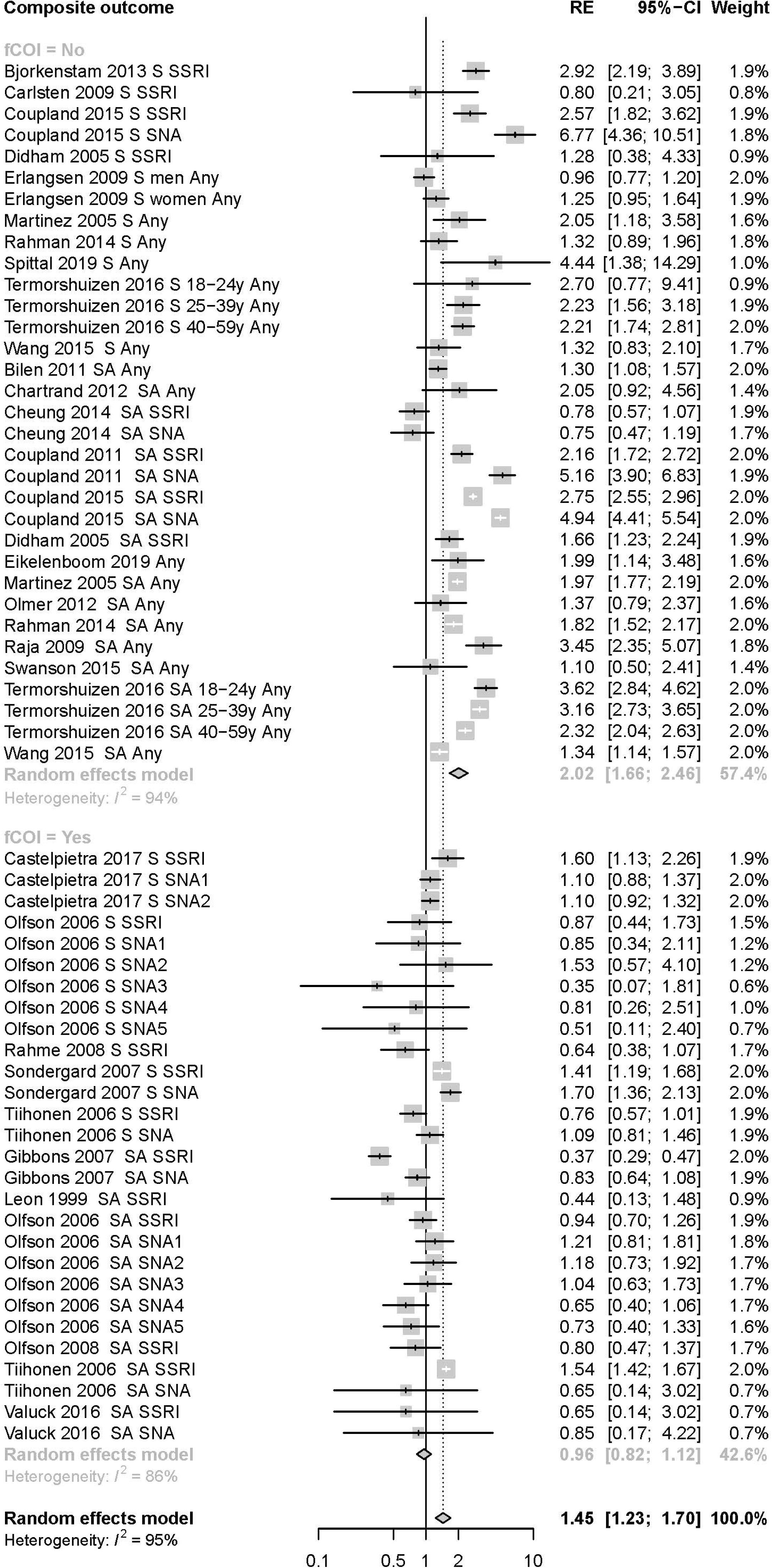

## Notes

### Competing Interest Statement

The authors have declared no competing interest.

### Clinical Protocols

https://osf.io/eaqwn/

### Funding Statement

No funding was received for this particular study

